# Effect of SARS-CoV-2 digital droplet RT-PCR assay sensitivity on COVID-19 wastewater based epidemiology

**DOI:** 10.1101/2022.04.17.22273949

**Authors:** Sooyeol Kim, Marlene K. Wolfe, Craig S. Criddle, Dorothea H. Duong, Vikram Chan-Herur, Bradley J. White, Alexandria B. Boehm

## Abstract

We developed and implemented a framework for examining how molecular assay sensitivity for a viral RNA genome target affects its utility for wastewater-based epidemiology. We applied this framework to digital droplet RT-PCR measurements of SARS-CoV-2 and Pepper Mild Mottle Virus genes made using 10 replicate wells, and determined how using fewer wells affected assay sensitivity and its performance for wastewater-based epidemiology applications. We used a computational, downsampling approach. When percent of positive droplets was between 0.024% and 0.5% (as was the case for SARS-CoV-2 genes during the Delta surge), measurements obtained with 3 or more wells were similar to those obtained using 10. When percent of positive droplets was less than 0.024%, then 6 or more wells were needed to obtain similar results as those obtained using 10 wells. When COVID-19 incidence is low, as it was before the Delta surge and SARS-CoV-2 gene concentrations are <10^4^ cp/g, using 6 wells will yield a detectable concentration 90% of the time. Overall, results support an adaptive approach where assay sensitivity is increased by running 6 or more wells during periods of low SARS-CoV-2 gene concentrations, and 3 or more wells during periods of high SARS-CoV-2 gene concentrations.

**Synopsis:** Adaptive approaches developed with assay sensitivity in consideration may reduce cost and increase sensitivity for wastewater-based epidemiology.

**Abstract Art:** 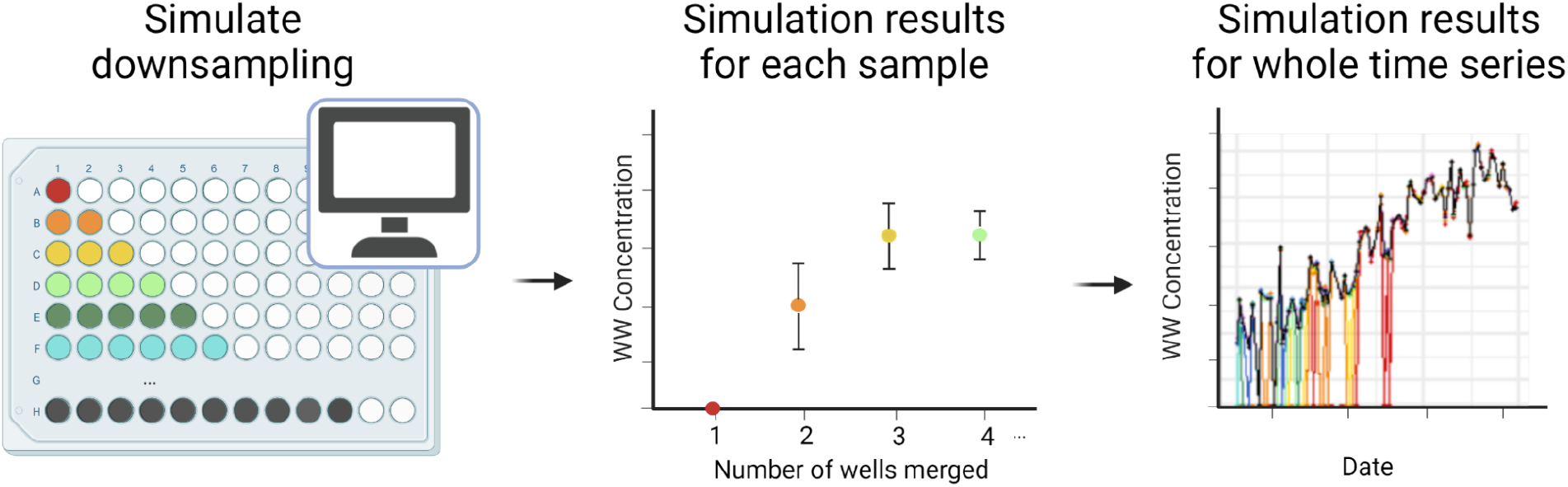

## Introduction

Wastewater-based epidemiology for severe acute respiratory syndrome coronavirus 2 (SARS-CoV-2) is becoming an increasingly important tool in monitoring coronavirus disease 2019 (COVID-19) incidence rates in communities. Monitoring programs that use samples from publicly owned treatment works (POTWs)^1,2^ and from sewer conveyances^3^ have actively been implemented to supplement clinical testing data. Wastewater surveillance has the advantage of providing insights into population health by overcoming limitations of clinical testing, such as test seeking behavior and test availability. Wastewater surveillance has also been used to gain insight into the epidemiology of other respiratory viruses such as influenza A virus^4^ and respiratory syncytial virus^5^ (RSV), as well as gastrointestinal pathogens such as hepatitis A^6^ and *Salmonella*^7^. Therefore, wastewater surveillance is likely to become increasingly useful for assessing various aspects of community health beyond COVID-19.

SARS-CoV-2 RNA concentrations in wastewater, whether measured in the solid or liquid phase or using quantitative or digital reverse-transcription polymerase chain reaction (RT-PCR), correlate to COVID-19 laboratory incidence rates in the population contributing to the wastewater^8–10^. The lower detection limit, or the sensitivity, of any method for detection of SARS-CoV-2 RNA, or any other disease target, will dictate the lowest levels of disease occurrence that can be detected using wastewater.

Digital (RT-)PCR can be a sensitive method for detecting disease targets in wastewater. Digital (RT-)PCR methods divide the entire (RT-)PCR solution (master mix, primers, probes and template) into a large number of partitions (droplets or physical partitions in a plate) such that each partition likely contains only one copy of template nucleic acid. By increasing the number of partitions (assuming their volume remains constant) and the associated reaction volume, the analytical sensitivity of the measurement can be increased. For digital droplet RT-PCR (ddRT-PCR), the number of partitions can be increased by increasing the number of wells used to analyze samples; the results from droplets generated by all replicate wells are merged to compute the final measurement. An analogous approach can be applied to other methods of digital partitioning. However, increasing the number of partitions used for each sample to improve sensitivity increases project reagent costs. In a previous study, we compared the lowest measurable concentration of SARS-CoV-2 RNA reported by different laboratories using different pre-analytical methods and ddRT-PCR, and the detection limit decreased as the number of replicate wells used in the method increased^11^. Authors have reported using between one to ten merged wells without full justification on how the number of wells merged was selected^2,11,12^.

Given the ongoing importance of wastewater surveillance for COVID-19 disease monitoring and its potential use for other disease surveillance, and increasing use of digital RT-PCR, it is important to better understand how analytical measurement sensitivity is controlled by increasing the number of partitions. To achieve this aim, we measured SARS-CoV-2 RNA and Pepper Mild Mottle Virus (PMMoV) RNA in wastewater solids samples using ddRT-PCR using 10 replicate wells. We then computationally down sampled the wells to investigate how the number of wells, and thus partitions, affects the lowest measurable concentrations of SARS-CoV-2 genes and associations between SARS-CoV-2 gene measurements and disease incidence rates. The framework developed herein for examining how molecular assay sensitivity for a viral RNA genome target affects its utility for wastewater-based epidemiology is generalizable to other infectious agents and other analytical approaches for measuring molecular targets.

## Materials and Methods

The Institutional Review Board of Stanford University determined that this project does not meet the definition of human subject research as defined in federal regulations 45 CFR 46.102 or 21 CFR 50.3 and indicated that no formal IRB review is required.

### POTWs and data collection

Data used in this study was obtained from an on-going SARS-CoV-2 wastewater monitoring program in California, USA described by Wolfe et al.^2^ Samples were collected and processed daily between June 1, 2021 to August 31, 2021 from four POTWs: City of Davis Wastewater Treatment Plant (Dav) in Davis, South County Regional Wastewater Authority Wastewater Treatment Plant (Gil) in Gilroy, Oceanside Water Pollution Control Plant (Ocean) in San Francisco, and San Jose-Santa Clara Regional Wastewater Facility (SJ) in San Jose (listed in the order of size from smallest to largest). Further details on the POTWs and sampling procedures can be found in Wolfe et al.^2^ and in Table S1. The data reported herein has not been previously published.

The solids samples were processed within 24 hours of collection exactly according to the methods described by Wolfe et al.^2^ and are summarized in the SI. We measured SARS-CoV-2 N gene and PMMoV gene concentrations, and also recovery of spiked-in bovine coronavirus using ddRT-PCR using 10 replicate wells. EMMI guidelines were followed^13^.

Data for each individual well was downloaded from QuantaSoft Analysis Pro software (BioRad, CA, version 1.0.596). Samples for which all wells did not have at least 10 000 generated droplets in each well were eliminated from our analysis. This eliminated a total of 41 SARS-CoV-2 N gene measurements (12 from Dav, 9 from Gil, 17 from Ocean, and 3 from SJ), resulting in 327 measurements for further analysis (89 from Dav, 80 from Gil, 83 from Ocean, 75 from SJ).

### COVID-19 epidemiology data

Laboratory confirmed incident cases of COVID-19 as a function of episode date was obtained as described previously^2^; see SI for details.

### Downsampling simulation

In order to estimate the SARS-CoV-2 N gene and PMMoV RNA concentration we would have obtained if we had run a smaller number of wells (X = 1 - 9), we randomly selected X wells from the 10 wells to calculate the resultant concentration:

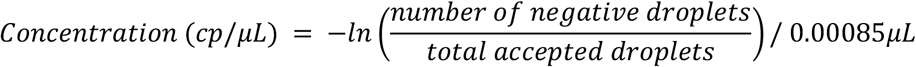

where 0.00085 μL is the volume of a single droplet^14^. If the total number of positive droplets was less than three, the concentration was denoted as not detected (ND).

A thousand simulations were conducted for each possible number of merged wells (X = 1 - 9) for each sample. The resulting concentrations were converted to units of cp/g dry weight using dimensional analysis ^2^. From these thousand simulations, we calculated (1) the percent of the simulations that resulted in less than 3 positive droplets across merged wells and was designated as ND, (2) the median concentration, and (3) its interquartile range (25th and 75th percentiles). To calculate the median, we substituted zero for ND. Similar analyses were performed for ten randomly selected PMMoV measurements from each POTW; each measurement had at least 10 000 total droplets generated in each well. Simulations were conducted using RStudio (version 1.4.1106).

### Statistical analysis

Statistics were computed using RStudio (version 1.4.1106). Shapiro-Wilk test was used to determine whether simulation outputs were normally distributed. The dispersion of the simulation outputs is defined by the interquartile range (IQR). The relative dispersion of the simulation outputs is described as the ratio of the median and the interquartile range. The dispersion and relative dispersion of the simulated concentrations were compared to the standard deviation of the concentration, or the standard deviation normalized by the concentration, respectively, derived from the measurement obtained using 10 wells. The standard deviation of that measurement is the 68% confidence interval as defined by the total error from the instrument software which includes Poisson error and variation among wells; the total error formula is proprietary and not available from the vendor.

Nonparametric Kendall’s tau was used to assess the association between the N gene concentrations in wastewater and laboratory confirmed COVID-19 incidence rates. Kendall’s tau was calculated for both the entire time series and the low incidence month of June. For this analysis, zero was substituted for measurements considered NDs.

The theoretical lower measurement limit was calculated for each number of merged wells (X = 1 - 10) by: 1) calculating the concentration resulting from three positive droplets total across merged wells out of 20 000 total accepted droplets (average number of droplets generated) per each well merged, and 2) converting the concentration to units of cp/g dry weight using average solid content for samples from each POTW (Table S2).

Linear regression was used to derive an empirical relationship between log_10_-transformed COVID-19 laboratory-confirmed incidence rates and log_10_-transformed measured SARS-CoV-2 N gene concentrations using data obtained by merging ten wells; relationships were quantified for each POTW separately, and for the POTWs in aggregate. For the linear regression, NDs were substituted with half the theoretical lower measurement limit calculated for X = 10. Using the empirical relationship between incidence rate and SARS-CoV-2 RNA concentration for the associated POTW, the lowest detectable COVID-19 incidence rate was estimated based on the calculated theoretical lower measurement limits.

A logistic regression was used to model the fraction of samples that were assigned a concentration (versus assigned ND) for X = 1 - 9 as a function of the true concentration of the sample, defined as the concentration obtained using 10 wells. The concentration corresponding to a detection frequency of 0.5 (C_0.5_) was calculated using the regression equation. For this analysis, half of the theoretical lower measurement limit was substituted for the 6 NDs for X =10.

## Results

### QA/QC

Negative and positive extraction and PCR controls were negative and positive, respectively. BCoV was used as a process control to verify that the extraction was successful and there was no gross inhibition in quantification. Samples that had less than 10% recovery of BCoV were rerun; no sample had less than 10% recovery. No further correction or analysis of BCoV recoveries are provided here given the complexities of interpreting recoveries of exogenous controls^15^. PMMoV concentrations across samples are similar to those measured and reported previously, also suggesting no gross issues with extraction or inhibition (Fig. S1). Data on N and PMMoV gene concentrations are available through the Stanford Digital Repository (https://purl.stanford.edu/km637ys9238).

### Simulation output trends at high and low concentrations

For each measurement, a thousand simulations were conducted to sample each possible number of merged wells (X = 1 - 9) and the results are reported as concentration in units of cp/g dry weight (cp/g, hereafter). The resulting concentration distributions obtained for each X for each measurement were not normally distributed based on Shapiro-Wilk tests (p < 0.05); therefore, medians and interquartile ranges are used to describe the results. Simulation outputs of example measurements for SARS-CoV-2 N gene in samples collected during a period of low COVID-19 incidence (June 1, 2021) and high COVID-19 incidence (August 31, 2021); as well as example PMMoV gene measurements (June 6, 2021) are provided in Figure 1. The simulation dispersion can be compared to the results obtained using 10 merged wells and its standard deviation, defined by the total error as reported by the ddPCR instrument, which includes errors associated with the Poisson distribution and variability among replicate wells.

**Figure 1.**
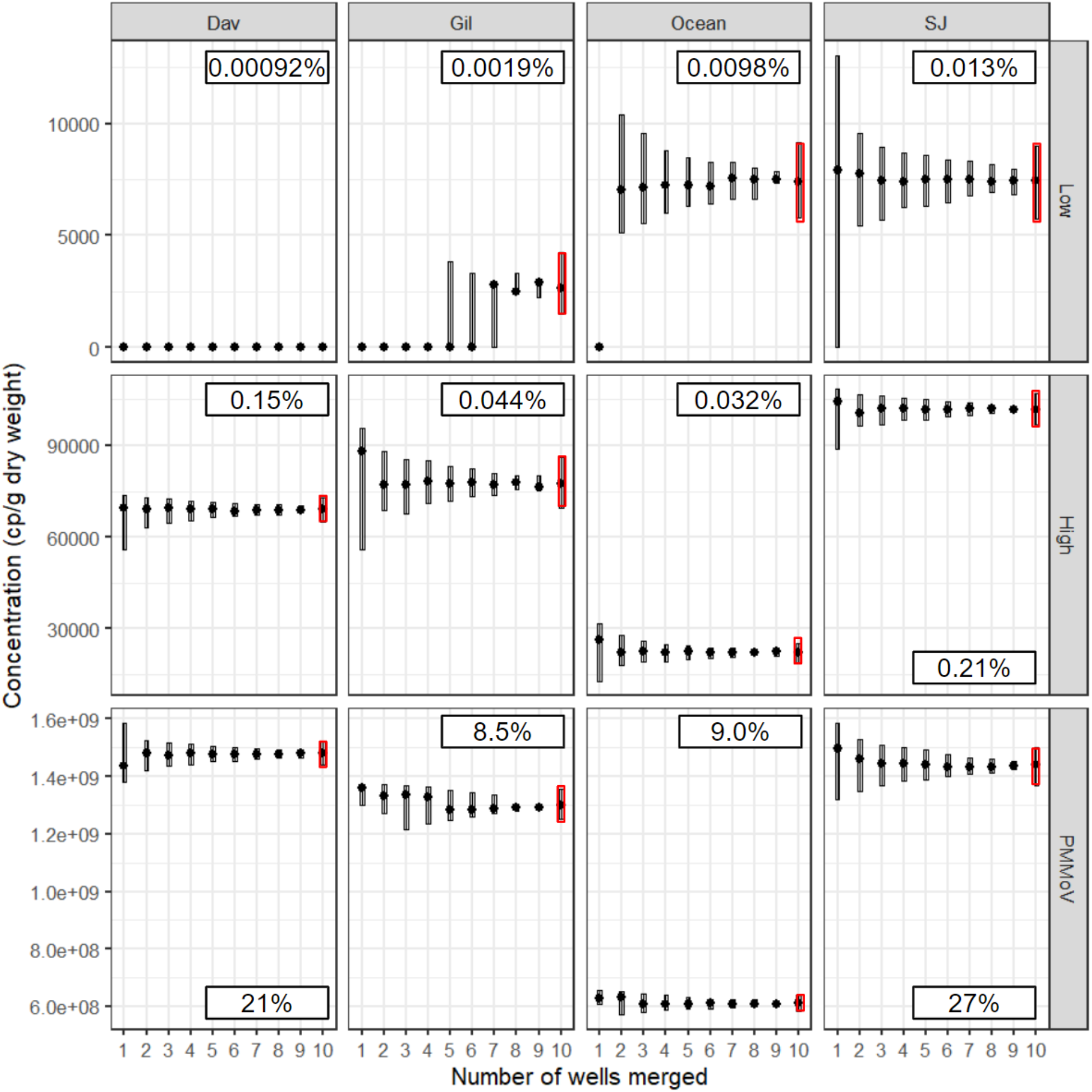
Example output of 1000 simulation results of selecting random sets of wells out of ten wells to calculate the final concentration in wastewater solids (cp/g dry weight) for SARS-CoV-2 N gene in June 1, 2021 sample during low COVID-19 incidence (top), SARS-CoV-2 N gene in August 31, 2021 sample during high COVID-19 incidence (middle), and for PMMoV in June 6, 2021 sample (bottom). For X = 1 - 9, the circle in the box represents the median, and the top and bottom of the box represent 75th and 25th percentile, respectively. For X = 10, the circle in the red box represents the software reported concentration from merging all ten wells, and the top and bottom of the box represent upper and lower confidence intervals, respectively, from 68% total error as given by the instrument software, which includes errors associated with the Poisson distribution and variability among replicate wells. Percentage of positive droplets in 10 wells is shown in boxes within each plot.

There are several important insights to glean from these results. First, when the number of positive droplets is less than 3 across the 10 wells (< 0.0015% of droplets positive), and the measurement is deemed as ND (see Dav sample from June 1, 2021), the results obtained from fewer wells agree with the results obtained from 10 wells. Second, when the number of positive droplets is high (for example, for PMMoV where there are 10^4^∼10^5^ positive droplets across 10 wells, 5 - 50% of droplets positive), then the dispersion as represented by the interquartile range (IQR), in the simulations for X < 10 wells is similar to the standard deviation reported by the instrument for X = 10 wells. Finally, when the number of positive droplets is intermediate to these two regimes (fraction of positive droplets between 0.0015% and 0.5%), then the IQR increases as X decreases and is often greater than the standard deviation from the X = 10 well measurement, particularly when X < 3. Below SARS-CoV-2 concentration of 10^4^ cp/g (where the fraction of positive droplets is 0.0062% - 0.024% depending on POTW), the value of X below which the relative dispersion, defined by IQR divided by the median, is larger than the standard deviation normalized by the measured concentration for X = 10 scales inversely with SARS-CoV-2 N gene concentration (Figure S2).

As PMMoV is present in such high concentrations and the measurements yielded high positive droplet counts, resulting in similar concentrations across X = 1 - 10, the remainder of this analysis will focus on the SARS-CoV-2 N gene concentrations, as those measurements yielded low to intermediate positive droplet counts.

### Theoretical sensitivity

An empirical relationship between the log_10_-transformed COVID-19 incidence rate and the log_10_- transformed SARS-CoV-2 concentration using ten merged wells was derived with linear regression. The regression showed that for 1 log_10_ increase in SARS-CoV-2 RNA cp/g, there was between 0.50 and 0.88 log_10_ increase in laboratory-confirmed COVID-19 incidence rate (Table S3), depending on the POTW. The data from all four POTWs appear to fall on a single line when COVID-19 incidence rate is plotted against SARS-CoV-2 concentration (Figure S3); when data from all POTW are combined, there was a 0.64 log_10_ increase in laboratory-confirmed COVID-19 incidence rate for 1 log_10_ increase in SARS-CoV-2 RNA cp/g.

Theoretical lower measurement limit (Table S4) and the corresponding incidence rate lower limit (Table S5) was calculated. The theoretical lower measurement limit for each POTW ranged from 7500 (SJ) to 24000 (Gil) cp/g when using only one well and from 750 (SJ) to 2400 (Gil) cp/g when using ten merged wells. Since this theoretical lower measurement limit was calculated with average solid content of samples from each POTW by measuring the percent weight of the dewatered solids and assuming a total of 20 000 generated droplets, the observed lower measurement limit may be different. The corresponding incidence rate lower limit per 100 000, calculated using the empirical relationships in Table S3, ranged from 1.6 (SJ) to 6.9 (Gil) when using one well and from 0.2 (SJ) to 1.4 (Gil) when using ten merged wells (Table S5).

### Association with clinical data

Time series of median concentrations resulting from a thousand simulations for each measurement for all possible numbers of wells are provided in Figure 2. Lines representing low number of wells deviate from those representing higher number of wells during the low incidence rate period in June 2021 when SARS-CoV-2 N gene concentrations in wastewater were relatively low. When the entire study period of June 1, 2021 to August 31, 2021 was considered, SARS-CoV-2 N gene concentrations were positively and significantly correlated with 7-day smoothed COVID-19 incidence rates at all four POTWs regardless of X (Table S6, tau > 0.54, p < 0.05 for all); X did not have an effect on Kendall’s tau when considering the entire time series (tau changed by < 0.05 as X varied). However, when considering the month of June alone when COVID-19 incidence rate was relatively low, SARS-CoV-2 N gene concentrations were positively and significantly correlated with 7-day smoothed COVID-19 incidence rates only when X > 1 (**α**= 0.1) and X did affect tau by as much as 0.15 (Table S7).

**Figure 2.**
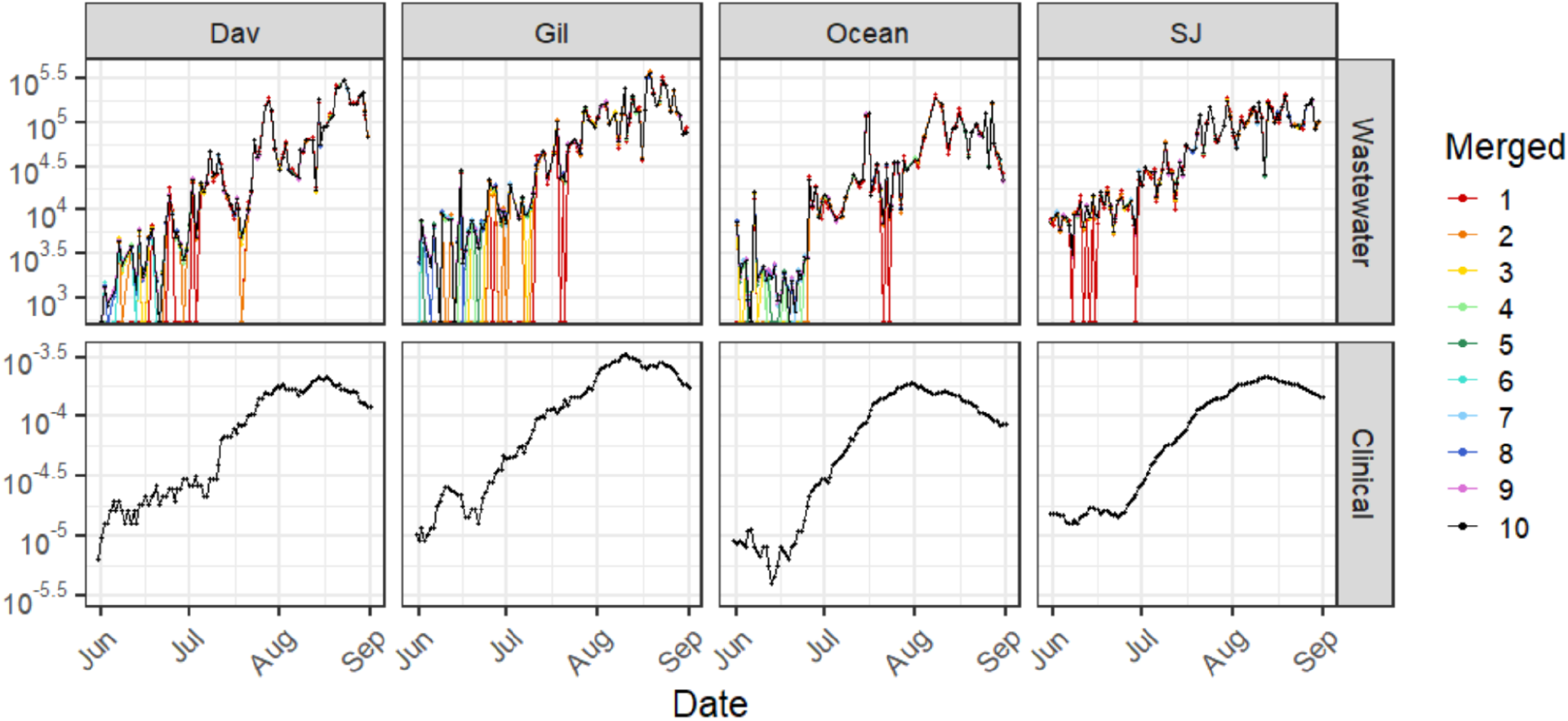
Time series of (top to bottom) SARS-CoV-2 N gene concentration in wastewater solids (cp/g dry weight) and 7 day centered smoothed average laboratory-confirmed SARS-CoV-2 incidence rate for each of the four POTWs from June 1, 2021 to August 31, 2021. Note that the SARS-CoV-2 N gene concentrations are displayed in log_10_-scale format for ease of visualization. Each wastewater data point for X = 1 - 9 represents median SARS-CoV-2 RNA concentration for a single sample obtained from 1000 simulations; for X = 10, each data point is the concentration obtained by merging 10 wells. Samples that resulted in ND were substituted with zero.

### Detection frequency

Detection frequency for SARS-CoV-2 N gene across all POTWs was examined as a function of X and the true concentration of N gene, as defined by the concentration obtained using 10 merged wells (Figure 3) Logistic regressions were fit to the curves (Table S8) and the concentration at which the detection frequency is ≤ 0.5 (C_0.5_) was calculated, as well as the corresponding incidence rate using the empirical relationship between log-transformed SARS-CoV-2 RNA N gene concentrations and COVID-19 incidence rate for all POTWs (Table 1). As there were not as many measurements that resulted in ND for X > 8, the confidence interval of the logistic regression is relatively large, especially for X = 9 and 10. Therefore C_0.5_ derived for X = 9 and10 is an extrapolation of the data used in this study and may be more uncertain. C_0.5_ scales with X according to the following equation log_10_(C_0.5_) = -0.13*X + 4.0 (R^2^ = 0.90, p-value < 10^−4^). Percentage of samples that fall under C_0.5_ for each X during the entire time series of three months and during the month of June was calculated. Most samples that fell under C_0.5_ were collected in June. Figure 4 shows data exclusively in June 2021 to focus on this time period; measurements obtained using X = 10 are shown with the concentration at which the detection frequency is ≤ 0.5 for X = 1,3, 6, 10 is shown in dashed lines. During this low incidence rate period, for X =1, 81% (92 of 113) measurements fall below the C_0.5_ across all POTWs. For X = 3, 38% (43 of 113) measurements, and for X = 6, 8.8% (10 of 113) fall below the corresponding C_0.5_. For reference, when X = 10, 4.4% (5 of 113) measurements fall below the C_0.5_ across all POTWs.

**Table 1.**
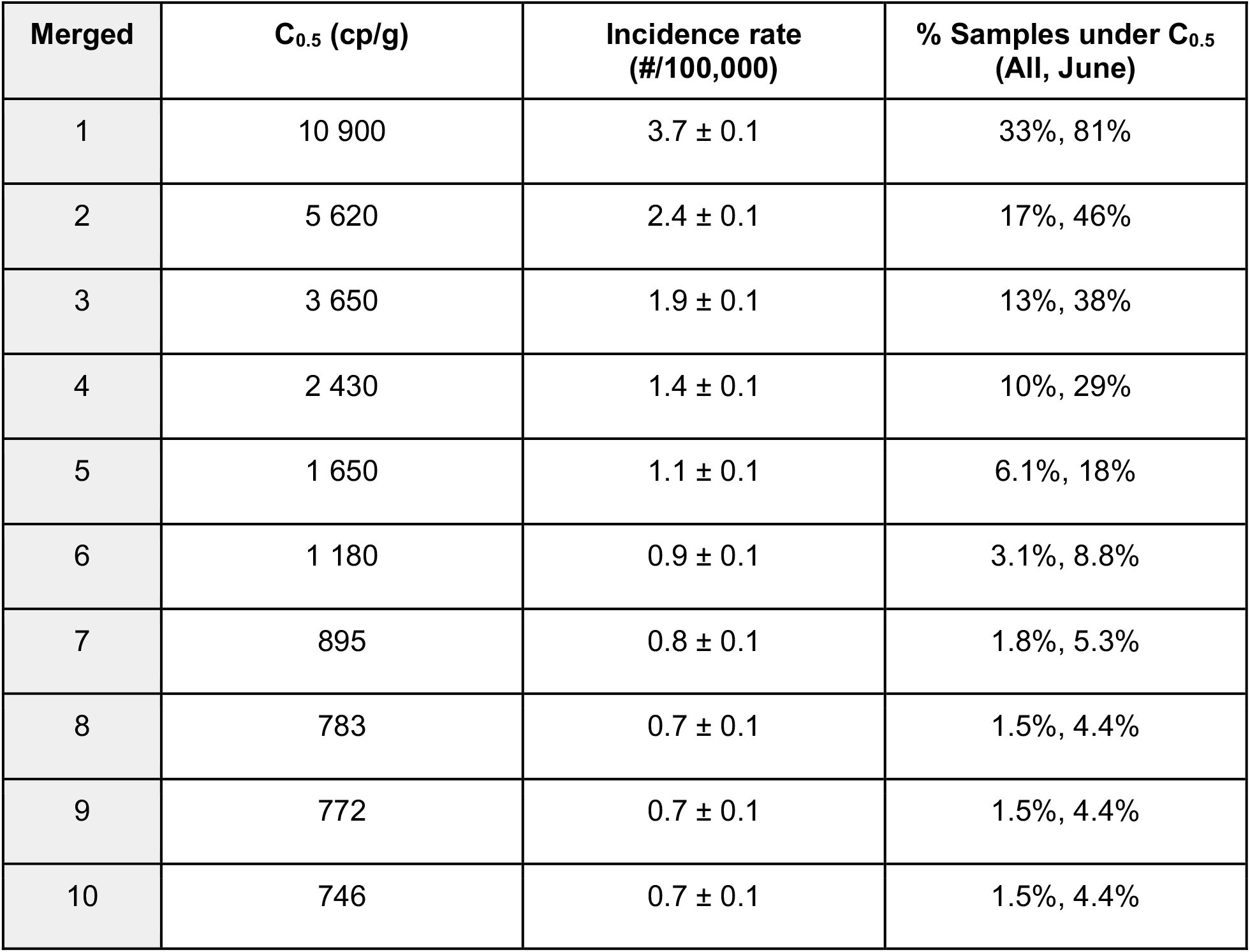
C_0.5_ shows the concentration that would yield a detect half the time in units of cp/g dry weight, calculated using the logistic regression (Figure 3). % Samples under C_0.5_ shows percentage of samples in the data set that were under C_0.5_ during the entire time series (All) and the month of June when COVID-19 incidence rate was low (June). Incidence rate shows the number of people out of 100,000 that corresponds to C_0.5_, calculated using the empirical relationship between log-transformed SARS-CoV-2 RNA N gene concentrations and laboratory-confirmed COVID-19 incidence rates using 10 merged wells for all POTWs derived in Table S3.

**Figure 3.**
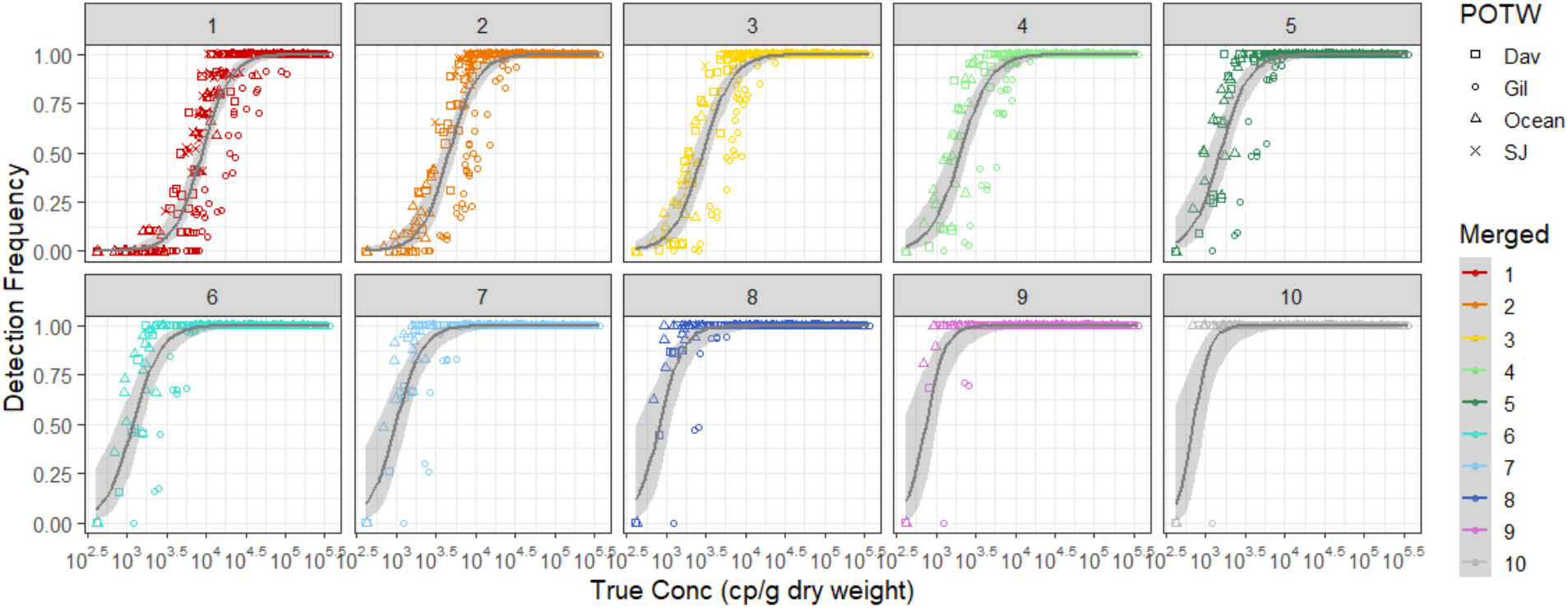
Detection frequency for all samples across four POTWs (y-axis) plotted against true concentration (x-axis), which is defined as concentration obtained by merging all ten wells. Each data point shows the fraction of 1000 simulations that did not result in ND in each well on the y- axis and its true concentration on the x-axis. ND for X = 10 was substituted with half of the theoretical lower measurement limit. 95% confidence intervals of the logistic regression are shown as the gray ribbon.

**Figure 4.**
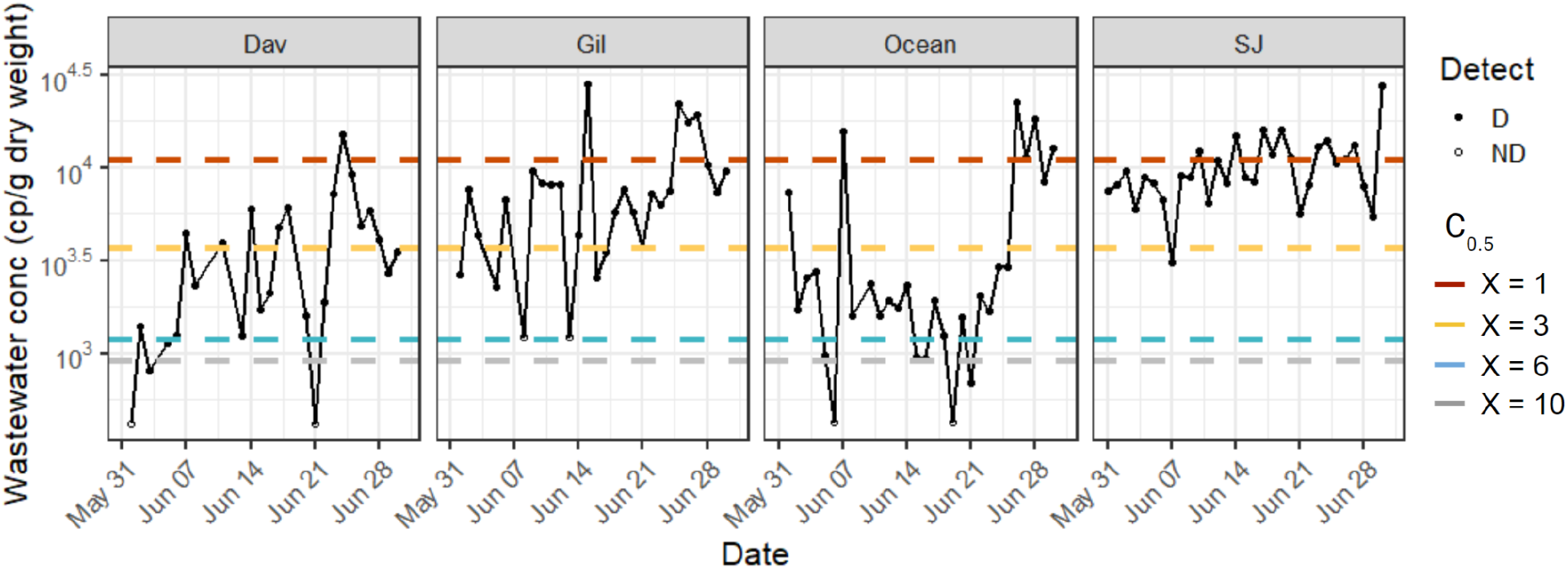
Time series of SARS-CoV-2 concentration in wastewater solids (cp/g dry weight) for each of the four POTWs during low COVID-19 incidence month of June 2021. Note that the SARS-CoV-2 RNA concentrations are displayed in log_10_-scale format for ease of visualization. Each wastewater data point represents SARS-CoV-2 concentration measured for a single sample. Samples above the lower measurement limit are shown as filled circles. Samples that resulted in ND, shown as empty circles, were substituted with half the lower measurement limit.

## Discussion

We measured concentrations of SARS-CoV-2 and PMMoV genes daily in wastewater settled solids at four POTWs in California using ddRT-PCR during a period of time that included both low and high COVID-19 incidence. We used 10 merged wells for the measurements, and then determined how the measurement would have been affected by using fewer than 10 wells through a down-sampling scheme. Our findings indicate that when a large fraction of droplets are positive (> 5% positive), as was observed for PMMoV, a virus found in high quantity in human stool and wastewater^16^, concentrations measured using just one well are similar to those obtained using ten when considering the variability associated with the measurements. On the other hand, when a smaller fraction of droplets are positive (< 0.5%), as was the case for the SARS-CoV-2 gene measurements, using fewer wells can result in measurements that may vary from those obtained using 10 wells, and more measurements characterized as non-detects.

For the SARS-CoV-2 gene measurements, variability in measurements increased as the number of wells decreased. Generally, we found that when the fraction of positive droplets was greater than 0.024% (corresponding to a conservative approximate concentration of 10^4^ cp/g dry weight), that the variability in the measurement resulting from using 3 or more wells was similar or smaller than the measurement total error obtained using 10 wells. In contrast, when the fraction of positive droplets was less than 0.024%, the variability in the measurements resulting from using 6 or more wells was similar or smaller than the measurement total error obtained using 10 wells. These results could guide adaptive analysis plans that use fewer wells to reduce costs when concentrations of SARS-CoV-2 are relatively high.

The probability of obtaining a non-detect increased as the SARS-CoV-2 gene concentration decreased and the number of wells used in ddRT-PCR decreased. Using logistic regression, we identified the concentration at which the detection frequency was less than 0.5 (C_0.5_), and this value varies inversely with the number of wells; that is C_0.5_ is higher when fewer wells are used for ddRT-PCR. This means that when low concentrations of SARS-CoV-2 genes are expected, using too few wells can result in a large number of non-detects. For example, during the low incidence period of June, if only 1 well had been used instead of 10, 92 of the 113 measurements across four POTWs would have been below C_0.5_. We found that at least 6 wells were needed to achieve 90% of the measurements to be above C_0.5_ for June 2021.

Consistent with other studies, the wastewater concentration showed positive and significant correlation with 7-day smoothed COVID-19 incidence rates. When there was variation in COVID-19 incidence rates within the time frame being investigated (here before and during the Delta variant surge), the number of wells being used for the analysis did not affect the magnitude or statistical significance of the correlation. There was a positive and significant correlation even when using only one well because there was enough variation in both variables, although the majority of June measurements were characterized as non-detects. This illustrates that finding a significant correlation between disease incidence and SARS-CoV-2 gene concentrations does not necessarily indicate good measurement sensitivity. It should be noted that while we take the laboratory confirmed COVID-19 incidence rates to be reflective of the level of COVID-19 that the community is experiencing, they are likely an underestimate of incidence rates in the sewershed as the reported incidence rates are dependent on test-seeking behavior and test availability^17^.

There are a few limitations of this analysis. First, in our analysis, we assumed that the measurement obtained using 10 wells is the “true concentration” and compared all results simulated with fewer than 10 wells to the true concentration and its error from the ddRT-PCR instrument. Second, the results presented herein regarding assay sensitivity, and in particular the C_0.5_ values in Table 1 are specific to the methods applied in this study. The relationship between the number of wells used to the number of non-detects, and the lowest measurable concentration will be impacted by the pre-analytical and analytical processes used. While the specific values in Table 1 are not externally valid, unless others are using our exact methods (available on protocols.io^18–20^), the framework for examining the required sensitivity for wastewater surveillance is. That is, careful attention to how sensitivity affects the lowest measurable concentration and the number of non-detects, as well as the relationships between these values and laboratory confirmed COVID-19 incidence rates is needed to fully understand how decisions on assay implementation are made.

## Conclusions

We developed and implemented a framework for examining how molecular assay sensitivity for a viral RNA genome target affects its utility for wastewater-based epidemiology. The framework involves understanding how assay sensitivity affects lowest measurable concentrations in units of copies per environmental matrix mass, and the detection probability of a target that is present; and how this change during periods of different disease occurrence can affect resultant statistical associations between the viral target and measures of disease incidence. We applied this framework to digital droplet RT-PCR (ddRT-PCR) measurements of a SARS-CoV-2 gene made using 10 replicate wells, and determined how using fewer wells affected assay sensitivity and its performance for wastewater-based epidemiology applications. From a reagent cost savings perspective, we recommend an adaptive analytical approach where assay sensitivity is increased by running more replicate wells (6 or more) during periods of low SARS-CoV-2 gene concentrations (using our methods, < 10^4^ cp/g) and COVID-19 incidence rate (< 3.5/100 000) and fewer replicate wells (3 or more) during periods of higher SARS-CoV-2 RNA concentrations and COVID-19 incidence. While the precise recommendations here are not generalizable, unless one is using the same pre-analytical and analytical protocols, the framework and the conclusion that adaptive approaches can reduce costs and increase sensitivity during periods of low disease incidence can be applied to other methods and other wastewater-based epidemiology targets.

## Data Availability

Data on N and PMMoV gene concentrations are available through the Stanford Digital Repository (https://purl.stanford.edu/km637ys9238).

## Supporting Information

Additional experimental, POTW (Table S1-S2), and COVID-19 epidemiological data details; simulation output (Table S3-S8); and results (Fig S1-S3).

## Acknowledgements

This study was supported by the CDC Foundation, NSF RAPID (CBET-2023057), and by the Epidemiology and Laboratory Capacity for Infectious Diseases Cooperative Agreement (no. 6NU50CK000539-03-02) from CDC. We thank the California Department of Public Health COVID-19 Wastewater Surveillance, Epidemiology and Data teams for their help with COVID-19 incidence data. We thank Dr. Linlin Li, Michael Balliet, Dr. Pamela Stoddard and Dr. George Han at the County of Santa Clara Public Health Department for provision of case data. Numerous people contributed to sample collection including including Payak Sarkar (SJ), Noel Enoki (SJ), Amy Wong (SJ), Alexandre Miot (Ocean), Lily Chan (Ocean), the Oceanside plant operations personnel, Saeid Vaziry (Gil), Chris Vasquez (Gil), and Jeromy Miller (Dav).

