## Supplemental Information for "Effect of SARS-CoV-2 digital droplet RT-PCR assay sensitivity on COVID-19 wastewater based epidemiology"

This supporting information consists of 8 pages including 8 tables and 3 figures.

### Brief description of experimental methods

Solids were dewatered and then a small mass was suspended in DNA/RNA Shield (Zymo Research, California) spiked with a known concentration of bovine coronavirus vaccine (BCoV, PBS Animal Health, Ohio, Calf-Guard Cattle Vaccine). The process of diluting the solid in the solution serves to alleviate inhibition[1]. The solution was then homogenized and then centrifuged. Subsequently, 10 replicate aliquots of the supernatant were subjected to nucleic acid extraction and inhibitor removal using commercial kits. The nucleic acids were used as a template in ddRT-PCR to measure the N gene of SARS-CoV-2, with each extraction replicate in its own well, consisting of a total of ten replicate wells for each sample. A 1:100 dilution of RNA extract was used as a template to measure BCoV, the spiked-in internal recovery control, and PMMoV, a fecal strength indicator and an endogenous internal recovery control, also run in 10 replicate wells. Extraction negative controls, extraction positive controls, no-template controls (NTC), and PCR positive controls were run on each plate. PCR positive controls consisted of guide RNA (gRNA) of N gene of SARS-CoV-2 (ATCC VR-1986D) and double-stranded DNA gene blocks for BCoV and PMMoV (Integrated DNA Technologies, Iowa).

### Details on COVID-19 epidemiology data

Earliest of reported symptom onset, laboratory result, or case record create dates for each sewershed was obtained from local or state sources. Case counts were georeferenced with residential addresses within the POTW service area. Incidence rate was calculated using the estimated population served by each POTW and a 7-day centered moving average was used in subsequent analysis.

Table S1. Details on publicly owned treatment work (POTW) and their sampling procedures: average daily inflow in million gallons per day (MGD), population served by the sewershed, and solid collection sampling point is shown.

| POTW | Average Daily Inflow (MGD) | Population Served | Solid Collection |
|---|---|---|---|
| Dav | 7.5 | 66 600 | Primary Clarifier |
| Gil | 8.5 | 110 300 | Settled solids collected from 24 hour composite raw influent using an Imhoff cone[2] |
| Ocean | 43 | 250 000 | Primary Clarifier |
| SJ | 167 | 1 458 000 | Primary Clarifier |

Table S2. Summary of fraction of solid measured for samples from each POTW; the average solid content was used to calculate the theoretical lower measurement limit.

| POTW | Dav | Gil | Ocean | SJ |
|---|---|---|---|---|
| Min | 0.108 | 0.048 | 0.147 | 0.171 |
| Average | 0.228 | 0.072 | 0.221 | 0.250 |
| Max | 0.32 | 0.111 | 0.313 | 0.321 |

Table S3. Empirical relationship between log-transformed SARS-CoV-2 RNA N gene concentrations and laboratory-confirmed COVID-19 incidence rates using 10 merged wells; the error represents standard error for the calculated coefficients.

| **POTW** | **Intercept** | **Slope** | **$R^2$** | **p-Value** |
|---|---|---|---|---|
| Dav | -6.32 ± 0.16 | 0.50 ± 0.04 | 0.70 | $< 10^{-15}$ |
| Gil | -7.26 ± 0.16 | 0.71 ± 0.03 | 0.84 | $< 10^{-15}$ |
| Ocean | -7.11 ± 0.20 | 0.67 ± 0.05 | 0.72 | $< 10^{-15}$ |
| SJ | -8.24 ± 0.16 | 0.88 ± 0.04 | 0.88 | $< 10^{-15}$ |
| All | -7.02 ± 0.09 | 0.64 ± 0.02 | 0.75 | $< 10^{-15}$ |

Table S4. Theoretical lower measurement limit (units of cp/g dry weight) for each POTW assuming the percent dry weight of dewatered solids was the average observed, 20 000 droplets generated per well for dd-RT-PCR, and 3 positive droplets across the merged wells.

| **Merged** | **Dav** | **Gil** | **Ocean** | **SJ** |
|---|---|---|---|---|
| 1 | 8200 | 24000 | 8500 | 7500 |
| 2 | 4100 | 12000 | 4300 | 3800 |
| 3 | 2700 | 8100 | 2800 | 2500 |
| 4 | 2100 | 6100 | 2100 | 1900 |
| 5 | 1600 | 4900 | 1700 | 1500 |
| 6 | 1400 | 4100 | 1400 | 1300 |
| 7 | 1200 | 3500 | 1200 | 1100 |
| 8 | 1000 | 3000 | 1100 | 940 |
| 9 | 920 | 2700 | 950 | 840 |
| 10 | 820 | 2400 | 850 | 750 |

Table S5. Measurable incidence lower limit calculated using empirical relationship derived from Table S3 for each number of merged wells and range of observed incidence rate calculated using theoretical lowest measurable concentration in Table S4 for each number of wells; the error is calculated from the standard error of the fit.

| | Merged | Dav | Gil | Ocean | SJ |
|---|---|---|---|---|---|
| Incidence Lower Limit (#/100 000) | 1 | 4.1 ± 0.3 | 6.9 ± 0.4 | 3.3 ± 0.3 | 1.6 ± 0.1 |
| | 2 | 2.9 ± 0.2 | 4.2 ± 0.3 | 2.1 ± 0.2 | 0.8 ± 0.1 |
| | 3 | 2.4 ± 0.2 | 3.2 ± 0.2 | 1.6 ± 0.2 | 0.6 ± 0.1 |
| | 4 | 2.1 ± 0.2 | 2.6 ± 0.2 | 1.3 ± 0.2 | 0.5 ± 0.1 |
| | 5 | 1.9 ± 0.2 | 2.2 ± 0.2 | 1.1 ± 0.1 | 0.4 ± 0.05 |
| | 6 | 1.7 ± 0.2 | 1.9 ± 0.2 | 1.0 ± 0.1 | 0.3 ± 0.04 |
| | 7 | 1.6 ± 0.2 | 1.7 ± 0.2 | 0.9 ± 0.1 | 0.3 ± 0.04 |
| | 8 | 1.5 ± 0.2 | 1.6 ± 0.2 | 0.8 ± 0.1 | 0.2 ± 0.04 |
| | 9 | 1.4 ± 0.2 | 1.5 ± 0.1 | 0.8 ± 0.1 | 0.2 ± 0.03 |
| | 10 | 1.3 ± 0.2 | 1.4 ± 0.1 | 0.7 ± 0.1 | 0.2 ± 0.03 |
| Range of observed incidence rate (#/100 000) | | 0 - 22 | 0.8 - 33 | 0.4 - 19 | 1.3 - 21 |

Table S6. Kendall's tau correlation between wastewater SARS-CoV-2 concentration and incidence rate in each sewershed for the entire study period between June 1, 2021 and August 31, 2021.

| | **Dav** | | **Gil** | | **Ocean** | | **SJ** | |
|---|---|---|---|---|---|---|---|---|
| | **Tau** | **p-Value** | **Tau** | **p-Value** | **Tau** | **p-Value** | **Tau** | **p-Value** |
| **1** | 0.62 | $< 10^{-14}$ | 0.70 | $< 10^{-17}$ | 0.57 | $< 10^{-11}$ | 0.69 | $< 10^{-20}$ |
| **2** | 0.62 | $< 10^{-14}$ | 0.72 | $< 10^{-19}$ | 0.57 | $< 10^{-11}$ | 0.70 | $< 10^{-21}$ |
| **3** | 0.63 | $< 10^{-15}$ | 0.73 | $< 10^{-21}$ | 0.58 | $< 10^{-12}$ | 0.70 | $< 10^{-21}$ |
| **4** | 0.63 | $< 10^{-15}$ | 0.74 | $< 10^{-22}$ | 0.57 | $< 10^{-12}$ | 0.70 | $< 10^{-21}$ |
| **5** | 0.63 | $< 10^{-15}$ | 0.74 | $< 10^{-22}$ | 0.57 | $< 10^{-12}$ | 0.70 | $< 10^{-21}$ |
| **6** | 0.63 | $< 10^{-15}$ | 0.74 | $< 10^{-22}$ | 0.56 | $< 10^{-11}$ | 0.70 | $< 10^{-21}$ |
| **7** | 0.63 | $< 10^{-15}$ | 0.74 | $< 10^{-22}$ | 0.55 | $< 10^{-11}$ | 0.70 | $< 10^{-21}$ |
| **8** | 0.63 | $< 10^{-15}$ | 0.74 | $< 10^{-21}$ | 0.55 | $< 10^{-11}$ | 0.70 | $< 10^{-21}$ |
| **9** | 0.63 | $< 10^{-15}$ | 0.73 | $< 10^{-21}$ | 0.55 | $< 10^{-11}$ | 0.70 | $< 10^{-21}$ |
| **10** | 0.63 | $< 10^{-15}$ | 0.73 | $< 10^{-21}$ | 0.56 | $< 10^{-11}$ | 0.70 | $< 10^{-21}$ |

Table S7. Kendall's tau correlation between wastewater SARS-CoV-2 concentration and incidence rate in each sewershed during the low incidence month of June.

| | **Dav** | | **Gil** | | **Ocean** | | **SJ** | |
|---|---|---|---|---|---|---|---|---|
| | **Tau** | **p-Value** | **Tau** | **p-Value** | **Tau** | **p-Value** | **Tau** | **p-Value** |
| **1** | 0.29 | 0.10 | 0.23 | 0.15 | 0.43 | $< 10^{-2}$ | 0.13 | 0.32 |
| **2** | 0.31 | 0.06 | 0.40 | $< 10^{-2}$ | 0.44 | $< 10^{-2}$ | 0.25 | 0.05 |
| **3** | 0.37 | 0.02 | 0.48 | $< 10^{-3}$ | 0.50 | $< 10^{-2}$ | 0.23 | 0.08 |
| **4** | 0.38 | 0.01 | 0.49 | $< 10^{-3}$ | 0.46 | $< 10^{-2}$ | 0.26 | 0.04 |
| **5** | 0.36 | 0.02 | 0.50 | $< 10^{-3}$ | 0.48 | $< 10^{-3}$ | 0.25 | 0.05 |
| **6** | 0.36 | 0.02 | 0.48 | $< 10^{-3}$ | 0.37 | $< 10^{-2}$ | 0.24 | 0.06 |
| **7** | 0.38 | 0.01 | 0.48 | $< 10^{-3}$ | 0.34 | 0.01 | 0.24 | 0.06 |
| **8** | 0.38 | 0.01 | 0.44 | $< 10^{-2}$ | 0.34 | 0.01 | 0.24 | 0.06 |
| **9** | 0.38 | 0.01 | 0.43 | $< 10^{-2}$ | 0.35 | 0.01 | 0.25 | 0.05 |
| **10** | 0.39 | $< 10^{-2}$ | 0.44 | $< 10^{-2}$ | 0.36 | $< 10^{-2}$ | 0.24 | 0.06 |

Table S8. Logistic regression between true concentration and fraction detected out of 1000 simulations for each measurement.

| Merged | Intercept | Estimate | p-value |
|---|---|---|---|
| 1 | -2.10 | $1.93 \times 10^{-4}$ | $< 10^{-10}$ |
| 2 | -2.26 | $4.02 \times 10^{-4}$ | $< 10^{-9}$ |
| 3 | -1.85 | $5.07 \times 10^{-4}$ | $< 10^{-8}$ |
| 4 | -1.47 | $6.04 \times 10^{-4}$ | $< 10^{-6}$ |
| 5 | -1.27 | $7.68 \times 10^{-4}$ | $< 10^{-4}$ |
| 6 | -1.13 | $9.59 \times 10^{-4}$ | $< 10^{-3}$ |
| 7 | -1.11 | $1.24 \times 10^{-3}$ | $< 10^{-2}$ |
| 8 | -1.41 | $1.80 \times 10^{-3}$ | $< 10^{-2}$ |
| 9 | -2.33 | $3.02 \times 10^{-3}$ | $< 10^{-2}$ |
| 10 | -3.29 | $4.41 \times 10^{-3}$ | $< 10^{-2}$ |

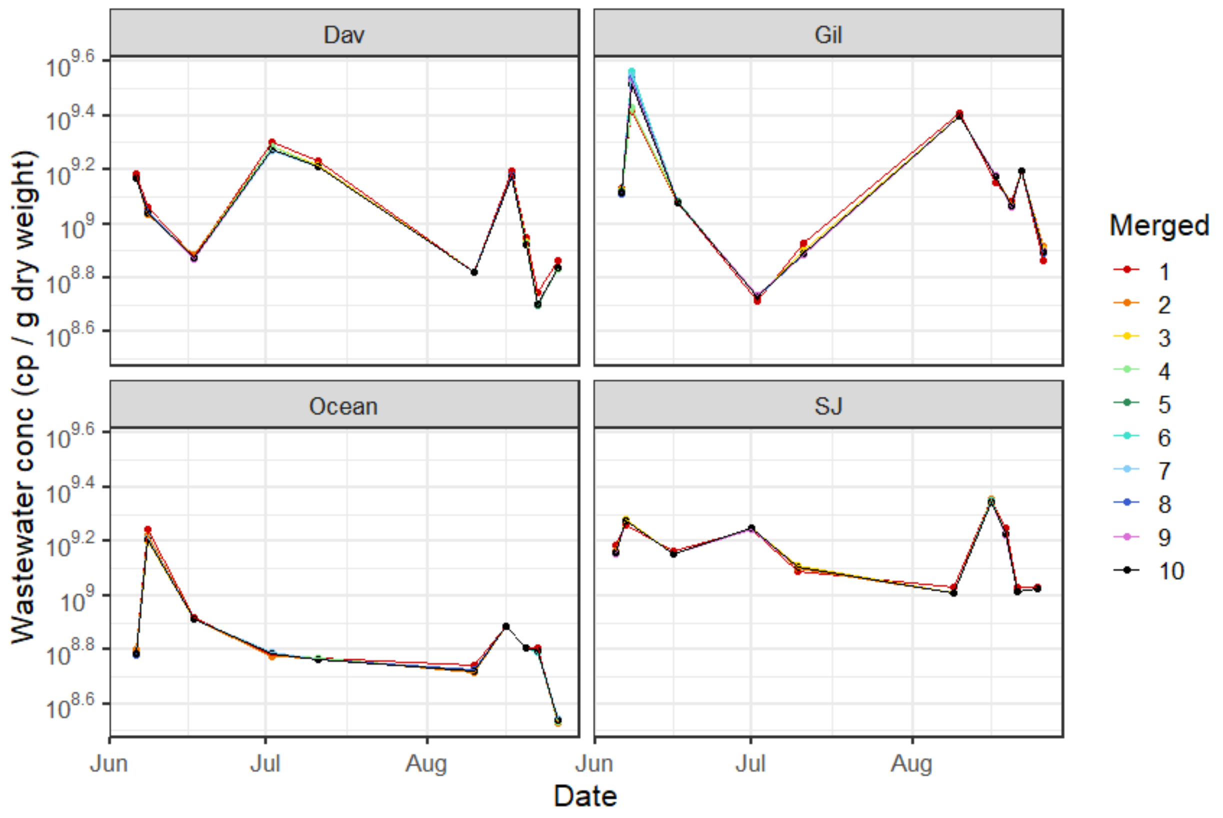


Figure S1. Time series of PMMoV concentration in wastewater solids (cp/g dry weight) from ten randomly selected samples for each of the four POTWs from June 1, 2021 to August 31, 2021.

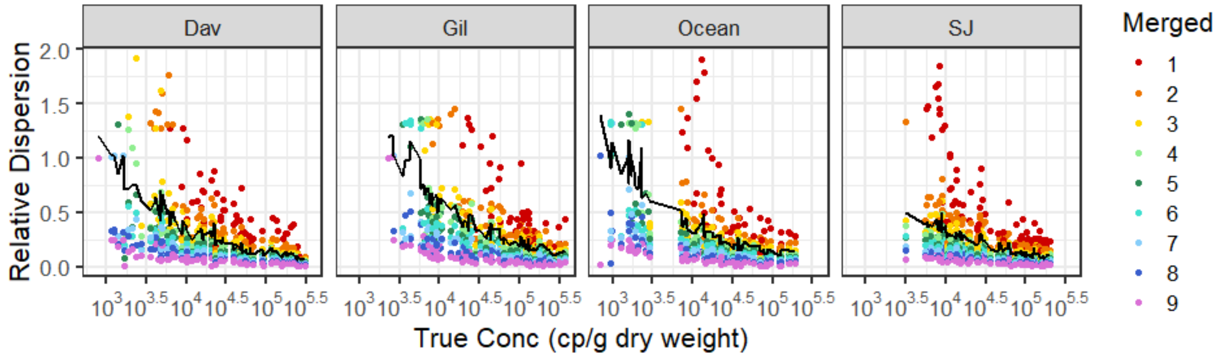


Figure S2. Relative dispersion (interquartile range normalized by median) of SARS-CoV-2 N gene concentrations resulting from thousand simulations for each number of merged wells plotted against true concentration, defined as concentration obtained from merging ten wells.

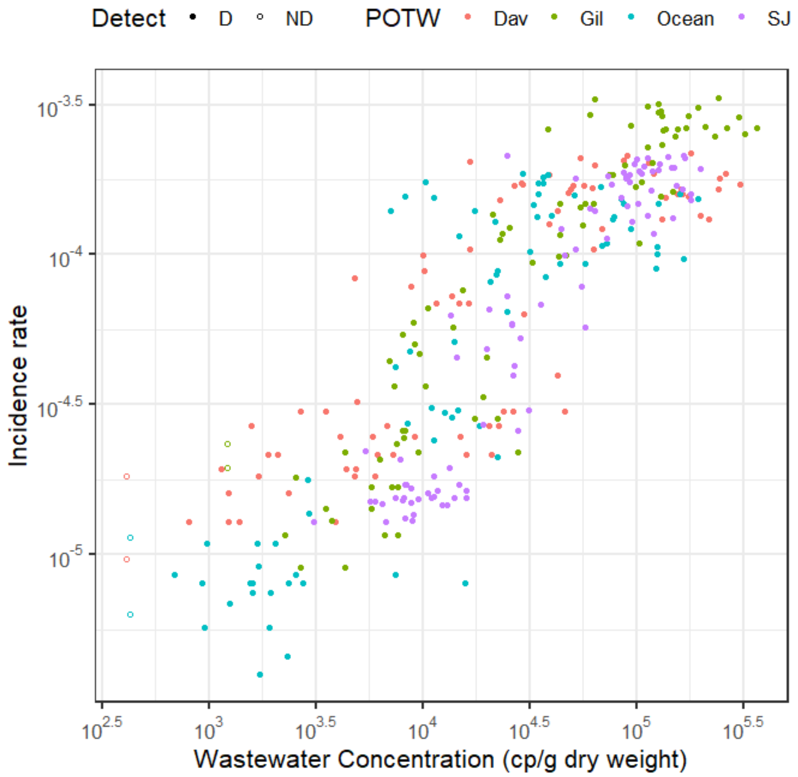


Figure S3. 7-day smoothed COVID-19 incidence rate plotted against SARS-CoV-2 concentration from merging ten wells.

(2) United States Environmental Protection Agency. Method 160.5, Methods and Guidance for the Analysis of Water Version 2.0, 1999.
